# Physical activity and health: Findings from Finnish monozygotic twin pairs discordant for physical activity

**DOI:** 10.1101/2021.11.11.21266038

**Authors:** Urho M. Kujala, Tuija Leskinen, Mirva Rottensteiner, Sari Aaltonen, Mika Ala-Korpela, Katja Waller, Jaakko Kaprio

**Author notes:** **Correspondence to:** Urho M. Kujala, Professor emeritus, Faculty of Sport and Health Sciences, P.O. Box 35, FI-40014 University of Jyväskylä, Finland.

## Abstract

Genetic and early environmental differences including early health habits associate with future health. To provide insight on the causal nature of these associations, monozygotic (MZ) twin pairs discordant for health habits provide an interesting natural experiment. Twin pairs discordant for leisure-time physical activity (LTPA) in early adult life are thus a powerful study design to investigate the associations between long-term LTPA and indicators of health and wellbeing. We have used this study design by identifying 17 LTPA discordant twin pairs from two Finnish twin cohorts and summarize key findings of these studies in this paper. The carefully characterized rare long-term LTPA discordant MZ twin pairs have participated in multi-dimensional clinical examinations. The occurrence of type 2 diabetes and death has been evaluated on the basis of prospective questionnaire data and register follow-up among a larger number of twin pairs. Key findings highlight that, compared to less active twins in such MZ twin pairs, the twins with higher long-term LTPA have higher physical fitness, reduced body fat, reduced visceral fat, reduced liver fat, increased lumen diameters of conduit arteries to the lower limbs, increased bone mineral density in loaded bone areas, increased high-density lipoprotein cholesterol levels and reduced occurrence of type 2 diabetes. However, we have not been able to document differences in the life expectancy between the less and more active twin siblings of the LTPA discordant MZ twin pairs. The findings are in agreement with intervention studies but not with all observational studies in which genetic factors are not controlled for.

## Introduction

Participation in leisure-time physical activity (LTPA) has been shown to be associated with many indicators of good health.^1^ However, based on observational studies, it is difficult to confirm the causality as many potential confounding factors including the genetic background influence these associations.^2^ Further, in short-term studies, reverse causality can bias results. It is possible that some genes may via the same mechanisms influence participation in physical activity, physical fitness, body composition, metabolism, and occurrence of chronic diseases (genetic pleiotropy).^3, 4^ Also, it is difficult to carry out high quality randomized controlled physical activity/exercise trials with long durations. Currently, there are no randomized controlled trials (RCTs) with the main outcome of occurrence of cardiac disease or mortality showing that increasing physical activity during adulthood is preventive. Trials with proxy outcomes can provide only suggestive but not definitive evidence.

The genetic background of physical activity, physical fitness and most of the non-communicable chronic diseases are multifactorial. Monozygotic (MZ) twin pairs are usually reared together in a similar environment until early adulthood. As genetic and early environmental differences including early health habits may have an influence on future health, MZ twin pairs who become discordance for LTPA during adult life provide an interesting natural experiment to investigate the associations between LTPA and indicators of health and wellbeing. Also, twins in MZ twin pairs who are discordant for physical activity usually are concordant for many other health habits.^5^ We have used this study design by comprehensively identifying LTPA discordant twin pairs from two Finnish twin cohorts (FinnTwin16 and older Finnish Twin Cohort). We summarize key findings of these studies in this paper. The carefully characterized rare long-term LTPA discordant MZ twin pairs have participated in multi-dimensional clinical examinations. In addition, the occurrence of type 2 diabetes and death has been evaluated on the basis of prospective questionnaire data and register follow-ups in the older Finnish twin cohort using a higher number of LTPA discordant pairs identified using 1 or 2 baseline questionnaires.

## Identification of the MZ twin pairs discordant for LTPA

The participants described here are from the Finnish population-based twin cohorts: FinnTwin 16 Cohort^6^ and the older Finnish Twin Cohort.^7^ The pairs were comprehensively selected from the cohorts with a stepwise process using available questionnaire data and subsequent detailed telephone and face-to-face interviews (Figure 1).^8, 9^ The twin pairs of the TWINACTIVE study (five male and two female MZ pairs) had been strongly discordant for LTPA for 30+ years and those of FITFATTWIN study (ten male MZ pairs) for 3 years. The MZ pairs were interviewed in depth using the same structural interview protocol for physical activity and data was pooled from the LTPA-discordant MZ pairs from both studies. The LTPA (including commuting activity) volume was 2.0±1.8 MET-hours per day for less active twins and 6.1±3.7 for their more active co-twins, the pairwise difference being 4.1 MET-hours per day (95% CI, 2.5 to 5.6; p<0.0001) in the pooled data (see Table 1 for selected pooled data and Supplemental Table 1 for selected data for TWINACTIVE and FITFATTWIN participants separately). The older TWINACTIVE pairs (mean age 62 years, range 50-74 years) represent pairs who were apparently healthy at baseline, but may have developed some predisease/disease states during follow-up prior to the clinical measurements. This may have an influence on the metabolic differences between the more and less active members of the MZ pairs in this older cohort. Among the younger FITFATTWIN participants (mean age 34 years, range 32-36 years) clinical disease was absent (and so does not have an influence on findings), thus difference in measured metabolic and clinical measures are showing more direct LTPA associations compared to those seen in older pairs. All the studies and their procedures that contributed to this summary review complied with the ethical standards of the relevant national and institutional committees on human experimentation and with the Helsinki Declaration of 1975, as revised in 2008. All participating twins provided informed consent.

**Figure 1.**
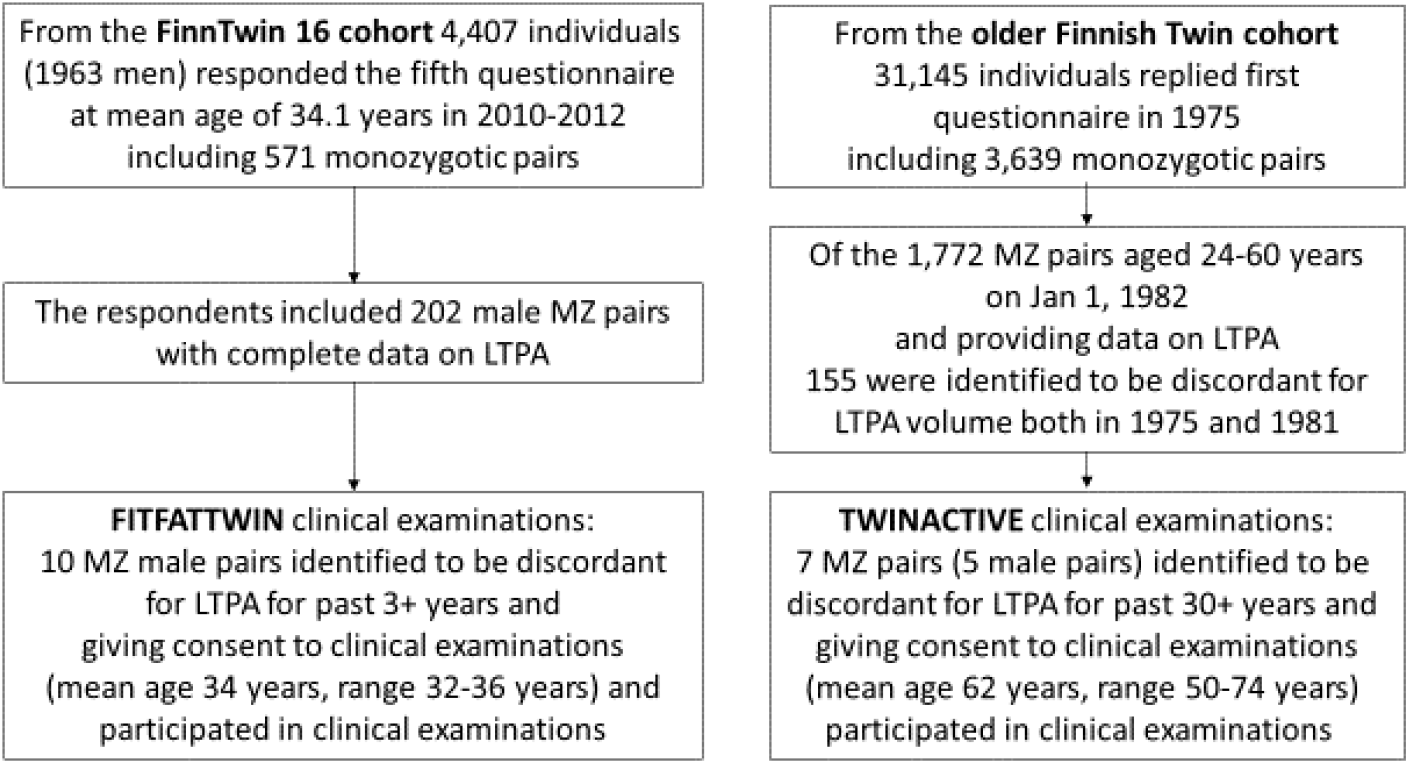
Identification of the rare leisure-time physical activity (LTPA) discordant monozygotic (MZ) twin pairs from the population-based twin cohorts.

**Figure 2.**
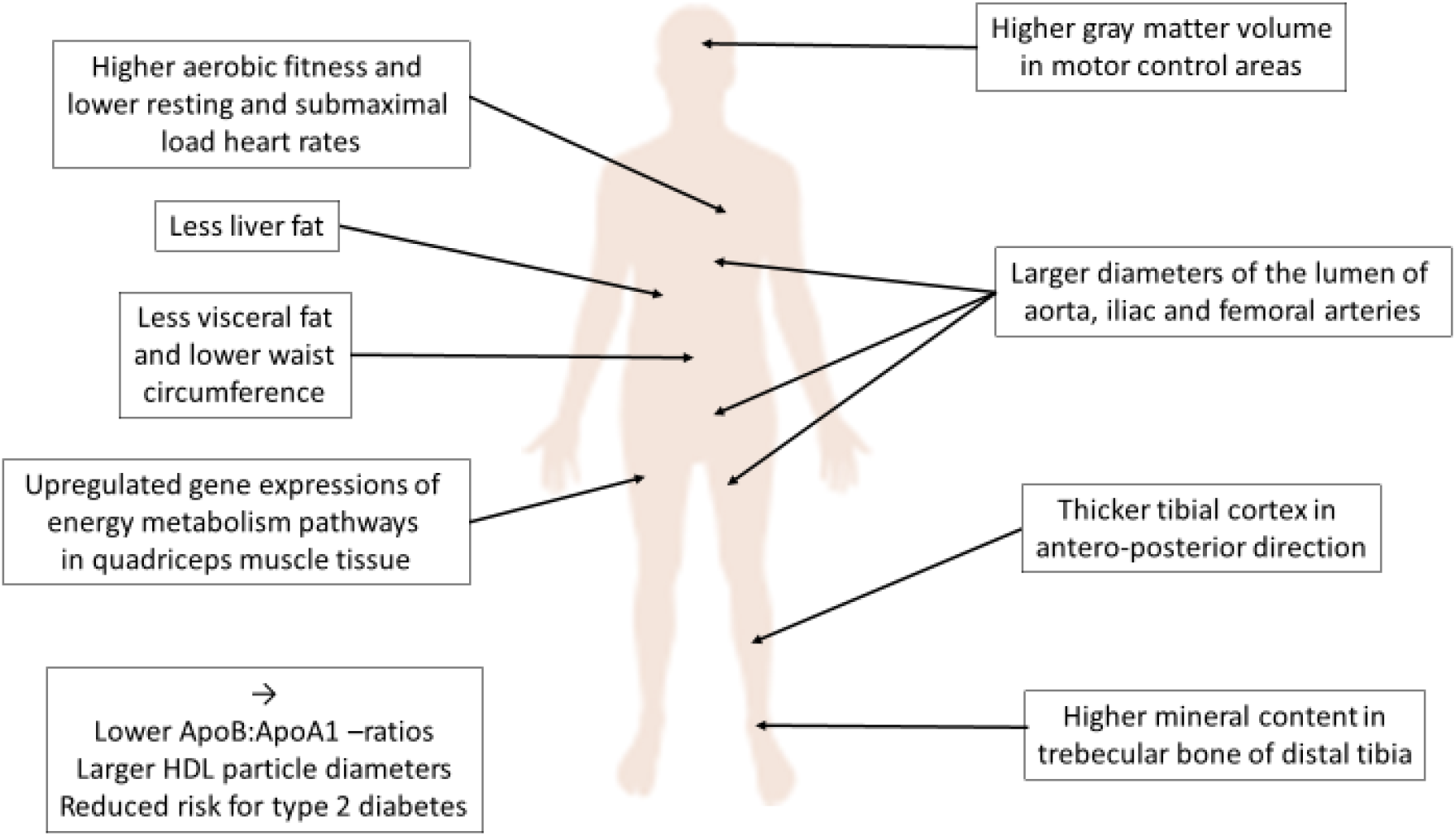
Key health-related findings showing how more physically active twins of monozygotic twin pairs differ from their less active co-twins.

**Table 1.** Selected data from monozygotic twin pairs discordant for leisure time physical activity habits pooled from TWINACTIVE and FITFATTWIN studies.

|  | Less active co-twins<br>(N=17) | More active co-twins<br>(N=17) | Intrapair difference<br>Mean (95% CI);<br>p-value |
| --- | --- | --- | --- |
|  | Mean±SD |  |  |
| VO<sub>2</sub>peak, ml/kg/min | 33.0±6.6 | 38.6±7.6 | 5.6 (3.3 to 8.0);<br>p=0.15x10<sup>-3</sup> |
| Past 12-month LTPA, LTMET-hours/day | 2.0±1.8 | 6.1±3.7 | 4.1 (2.5 to 5.6)<br>p=0.53x10<sup>-4</sup> |
| Waist circumference, cm | 92.3±9.0 | 88.4±7.8 | -3.9 (-6.9 to -0.8);<br>p=0.016 |
| Fat percent, % | 22.6±6.0 | 19.7±4.7 | -3.0 (-5.6 to -0.6);<br>p=0.027 |
| Visceral adipose tissue area, cm<sup>2</sup> | 144±54 | 107±55 | -37 (-61 to -13);<br>p=0.0045 |
| Liver fat index, MRI signal intensity | 15.1±16.6 | 6.8±5.9 | -8.3 (-15.4 to -1.1);<br>p=0.026 |
| ApoB:ApoA1 ratio | 0.607±0.133 | 0.529±0.034 | -0.079 (-0.133 to -0.024);<br>p=0.0077 |
| HDL cholesterol, mmol/L | 1.427±0.096 | 1.540±0.105 | 0.113 (0.011 to 0.216);<br>p=0.033 |
| HDL<sub>2</sub> cholesterol, mmol/L | 0.928±0.347 | 1.045±0.416 | 0.118 (0.017 to 0.218);<br>p=0.025 |
| HDL diameter, nm | 9.722±0.230 | 9.830±0.237 | 0.108 (0.054 to 0.162);<br>p=0.65x10<sup>-3</sup> |
| Very large HDL particles, mmol/L | 2.068x10<sup>-4</sup>±0.1334x10<sup>-4</sup> | 2.728x10<sup>-4</sup>±1.566x10<sup>-4</sup> | 0.660x10<sup>-4</sup><br>(0.171x10<sup>-4</sup> to 1.148x10<sup>-4</sup>);<br>p=0.011 |
| Large HDL particles, mmol/L | 8.018 x10<sup>-4</sup>±4.340 x10<sup>-4</sup> | 9.762x10<sup>-4</sup>±4.977x10<sup>-4</sup> | 1.744x10<sup>-4</sup><br>(0.769x10<sup>-4</sup> to 2.719x10<sup>-4</sup>);<br>p=0.0016 |
| Medium HDL particles, mmol/L | 18.197x10<sup>-4</sup>±4.667x10<sup>-4</sup> | 18.151x10<sup>-4</sup>±4.341x10<sup>-4</sup> | -0.045x10<sup>-4</sup><br>(-1.323x10<sup>-4</sup> to 1.232 x10<sup>-4</sup>);<br>p=0.94 |
| Small HDL particles, mmol/L | 46.383x10<sup>-4</sup>±3.564x10<sup>-4</sup> | 45.453x10<sup>-4</sup>±3.925 x10<sup>-4</sup> | -0.930x10<sup>-4</sup><br>(-2.491x10<sup>-4</sup> to 0.632x10<sup>-4</sup>);<br>p=0.23 |

Abbreviations: CI, confidence interval; LTMET-hours/day, leisure-time metabolic equivalent for physical activity during leisure-time and the commute to and from work indicating daily leisure-time physical activity volume; MRI, magnetic resonance imaging; ApoB, apolipoprotein B; ApoA1, apolipoprotein A1; HDL, high density lipoprotein.

## Reasons for the LTPA discordances

In the small number of LTPA discordant MZ pairs statistically significant reasons for the discordances could not be identified. However, it is to note that when motives for LTPA were measured in the larger twin cohorts, the motivational factors for LTPA that differed significantly between the more physically active twins and their less physically active co-twins in both the cohorts were mastery, physical fitness, and psychological state.^10, 11^ In young adulthood, family and work-related commitments obviously influenced the occurrence of LTPA discordances between the members of the twin pairs.^9^ Interestingly, however, the barriers to exercise training (including e.g. lack of time or lack of facilities for exercise) did not differ between the less and more active twins at an older age.^10^

## Main findings on the MZ pairs discordant for LTPA

The selection of the twin pairs to each substudy and their results have been previously published and the used measurement methods described in detail in the references given for each outcome. The pooled results given in Table 1 are previously unpublished.

### Fitness and heart function

According to pairwise analyses aerobic fitness (measured by a maximal exercise test using bicycle ergometer^8, 9^) was higher in the more active twins compared to their less active co-twins (for pooled results see Table 1 and for the results of younger and older pairs separately, see supplementary Table 1). The heart rate was lower in physically more active twins compared to their less active co-twins both at rest and during identical submaximal loads.^12^

### Body weight and fat accumulation

In both the FITFATTWIN and TWINACTIVE studies body weight, BMI and fat free mass did not differ between the members of the pairs as strongly as body fat composition.^13, 14^ When data was pooled from these two studies, among the 17 MZ pairs waist circumference, body fat percent, liver fat content, and in particular visceral fat was lower in active compared to inactive twins (Table 1). Given the clear differences in LTPA levels, the observation that dietary energy intake tended to be higher in more physically active members, indicates that different LTPA levels are the most plausible explanation for these body composition differences.^14, 15^

### Risk factors/metabolites measured from venous blood samples

Various associations in metabolites with high LTPA were seen in the TWINACTIVE study.^16^ In the pooled data from TWINACTIVE and FITFATTWIN studies, we saw that the more active twins had lower ApoB:ApoA1 –ratios (difference -0.079; 95% CI -0.133 to -0.024, p=0.0077), higher HDL2 cholesterol levels (difference 0.118 mmol/L; 95% CI 0.017 to 0.218, p=0.0247), and higher HDL particle diameters (difference 0.108 nm; 95% CI 0.054 to 0.162, p=0.0007) (Table 1) compared to the less active twins. The HDL particle size has been shown to be associated with its functions.^17^

### Muscle and adipose tissue gene expressions

In the TWINACTIVE study muscle biopies were obtained from the twin pairs. Gene expression in skeletal muscle of the central pathways of energy metabolism, especially of genes related to the processes of oxidative phosphorylation were up-regulated among the more physically active twins.^18^ Interestingly, the upregulation of skeletal muscle oxidative phosphorylation gene set expression correlated with the number of large HDL particles (r=0.75, p=0.0003), but not with the number of small HDL particles^18^ supporting the idea that the functions of HDL particles include associations with exercise related oxygen use in skeletal muscles. In both skeletal muscle and subcutaneous fat tissue samples, the up-regulated pathways among the active twins compared to their inactive co-twins included branched-chain amino acid (BCAA) degradation^18^ as an indicator of increased BCAA degradation in healthy mitochondria to uphold lipid oxidation. So, serum BCAA levels correlate with low LTPA and high body fat as has been shown earlier by Felig *et al*.^19^ and by Pietiläinen *et al*.^20^ among MZ twin pairs discordant for BMI.

### Arteries to lower limbs

The TWINACTIVE study used contrast-enhanced magnetic resonance angiography to measure the diameters of aorta, iliac and femoral arteries. Compared to less active co-twins, the active twins had larger lumen diameters in these lower limb arteries.^21^ A similar difference was not seen in the size of carotid arteries, suggesting location-specific effects.

### Tibial bones

The TWINACTIVE study using peripheral quantitative computer tomography showed that, compared to less active twins of these MZ twin pairs, the more active ones had thicker tibial cortical bone in antero-posterior direction and higher trabecular bone density in the distal tibia.^22^ There were no clear differences in the external dimensions of the bones which are likely to change more during growth. The documented changes were clearly site-specific according to the loading.

### Brain structure and functions

The FITFATTWIN study showed that total-brain white matter, grey matter (GM), and total intracranial volumes did not differ between the more and less active twins when analyzed with magnetic resonance imaging whole-brain voxel-based morphometry. However, compared to the less active twins, the more active co-twins showed larger striatal and non-dominant inferior frontal gyrus GM volumes.^9^ Also, there was regional differentiation in GM volumes between more and less active co-twins suggesting higher GM volume in the left hippocampus in more active co-twins.^23^ These findings provide evidence for site-specific structural modulation in healthy young adult brain likely associated with long-term LTPA. Additional comparisons showed differing automatic deviance-detection processes in brain regions involved with sensorimotor, visual and memory functions in electrophysiological studies between the more and less active co-twins.^24, 25^

## Type 2 diabetes and mortality in larger numbers of LTPA discordant MZ twin pairs

### Type 2 diabetes

In the older Finnish twin cohort type 2 diabetes occurred less often or later in the members of MZ twin pairs who had higher levels of LTPA compared to their less active co-twins^26^ supporting the metabolic health benefits of LTPA.

### Mortality

In the entire older Finnish twin cohort we have investigated how LTPA is associated with mortality using differing lengths of LTPA discordances and follow-ups. At baseline in 1975, 7925 healthy men and 7977 healthy women of the older Finnish Twin Cohort (aged 25 to 64 years) responded to a questionnaire on physical activity habits and known predictors of mortality.^27^ The results have shown that in the individual-based analyses high physical activity is associated with reduced risk of future death. In a follow-up study with more deaths, we observed that also in dizygotic (DZ) same-sex twin pairs the more active twin has reduced risk of death compared to the less active co-twins. No difference was seen for MZ pairs discordant for LTPA but the number of deaths in such pairs was modest.^28^ We further repeated the pairwise analyses among DZ and MZ pairs separately using cohort participants, who were discordant for vigorous physical activity in 1975 and 1981 with a follow-up until August 2013.^29^ This dataset included 778 DZ and 231 MZ vigorous LTPA discordant twin pairs. Among these DZ pairs there were 204 deaths and among MZ pairs 55 deaths during follow-up (between 1981 and 31^st^ August 2013). Unadjusted pairwise HR for death among active compared to inactive twins for DZ pairs was lower (HR 0.58 [95% CI 0.46–0.74]) than among MZ pairs (0.85 [0.56–1.30]). When this secondary analysis was repeated in the baseline-healthy subgroup of participants, the HRs were 0.64 (0.45–0.89) for DZ pairs and 1.05 (0.58–1.88) for MZ pairs, respectively. The heritability of physical activity is likely to contribute to the fact that there is low number of MZ pairs who are long-term discordant for physical activity and thus the analyses have only moderate statisical power. Moreover, no differences for the risk of death by work-related physical activity were observed in the pairwise analyses of all twin pairs.^29^ Our MOBILETWIN study which was a 40-year follow-up to a subgroup of the older Finnish Twin Cohort including accelometer measurements of physical activity^30^ showed that genetic factors contributed to the tracking of physical activity from middle age to old age. At older ages occurrence of diseases contributes to the physical activity discordances.^31^

More data on the findings is also available in nine PhD theses (see links to the texts in Supplement 2).

## Strengths and limitations of the co-twin control study design

As a result of the match for age, sex, gene sequence, and a close match for intrauterine and childhood environment, the MZ co-twin control study represents a unique study design to investigate the health effects of long-term physical activity controlling for genetic and familial factors. However, it is uncommon that co-twins of a MZ twin pair have persistently different activity levels, and it is therefore difficult to find large number of twin pairs significantly discordant for physical activity. This limitation itself speaks for a genetic and other familial basis for lifetime activity patterns. Consequently, the number of LTPA discordant MZ pairs remained small in both studies. We included extensive retrospective questionnaire-based follow-ups of LTPA habits combined with detailed structured interviews in our studies. Although the data relays on self-reported measures of physical activity, taking into account the limitations of device-based physical activity measurement methods and their non-existence over 40+ years ago, our data provide reliable estimates of long-term LTPA differences.^32^ It is to note that other health habits and socioeconomic status of the members of MZ pairs are more similar than between randomly selected individuals, and our twin pairs did not include those with strong discordance for smoking or alcohol use. Because the number of subjects in both studies remained quite low, we were able to carry out in depth clinical examinations, which usually are not included in large population studies.

## Comment

As many of the associations between LTPA and health outcomes were seen in the pairwise analyses among MZ twin pairs, our study findings support the existence of causal relationships. Expectedly, the pairwise differences were larger among the members of the older twin pairs with longer and stronger differences in LTPA, see supplement 1. The present data from LTPA discordant MZ twin pairs are in agreement with the data from RCTs^33^ showing that exercise inteventions improve fitness, body composition and selected cardiometabolic disease risk factors. Our results are also in line with intervention data on the lack of effect of adult-onset LTPA on mortality and thus not supporting the suggestive observational data from population cohorts. In the future, collaborative studies using different twin cohorts with higher number of LTPA discordant twin pairs are warranted to add to the current knowledge.

## Data Availability

All data produced in the present study are available upon reasonable request to the authors.

## Acknowledgements

We thank the participants of the reported twin studies and the collaborating authors of different specific research reports.

## Main funding of the substudies

TWINACTIVE study: The Finnish Ministry of Education and Culture (grant to UMK), Academy of Finland (grant to UMK), Finnish Cultural Foundation (grant to TL), Juho Vainio Foundation (grants to UMK, TL and SA) and Yrjö Jahnnsson Foundation (grant to SA).

FITFATTWIN study: The Finnish Ministry of Education and Culture (UMK), META-PREDICT (within the European Union Seventh Framework Program, HEALTH-F2-2012-277936 to UMK), Juho Vainio Foundation (MR), and Finnish Cultural Foundation (MR).

MOBILETWIN study: The Finnish Ministry of Education and Culture (grant OKM/56/626/2013 to UMK).

The Finnish Twin Cohort Study has been supported by multiple grants over the past 45 years, including grants from the Academy of Finland (to JK) and NIH (to JK and Richard J Rose for FinnTwin16). The current grant (# 336823) from the Academy of Finland supporting JK is specifically acknowledged. MAK is supported by a research grant from the Sigrid Juselius Foundation, Finland.

## Supplementary information

Supplementary information includes Supplementary Table 1 with selected data from monozygotic twin pairs discordant for leisure time physical activity habits, TWINACTIVE and FITFATTWIN studies, and Supplement 2 with links to PhD theses (Sports and Exercise Medicine, University of Jyväskylä) which include results from leisure-time physical activity discordant twin pairs.

## References

1. 2018 Physical Activity Guidelines Advisory Committee. 2018 Physical Activity Guidelines Advisory Committee Scientific Report. Washington, DC: U.S. Department of Health and Human Services, 2018.

2. Kujala UM. Is physical activity a cause of longevity? It’s not as straightforward as some would believe. A critical analysis. BJSM. 2018;52:914–8.

3. Kujala U. Physical activity, genes, and lifetime predisposition to chronic disease. Eur Rev Aging Phys Act. 2011;8:31–6.

4. Sillanpää E, Palviainen T, Ripatti S, Kujala UM, Kaprio J. Polygenic score for physical activity is associated with multiple common diseases. Med Sci Sports Exerc. 2021; online ahead of print: doi: 10.1249/MSS.0000000000002788.

5. Kaprio J, Koskenvuo M, Sarna S. Cigarette smoking, use of alcohol, and leisure-time physical activity among same-sexed adult male twins. Prog Clin Biol Res. 1981;69 Pt C:37–46.

6. Kaidesoja M, Aaltonen S, Bogl L, Heikkilä K, Kaartinen S, et al. FinnTwin 16: A longitudinal study from age 16 of a population-based Finnish twin cohort. Twin Res Hum Genetics. 2019;22:530–9.

7. Kaprio J, Bollepalli S, Buchwald J, Iso-Markku P Korhonen T, Kovanen V, et al. The older Finnish Twin Cohort – 45 years of follow-up. Twin Res Human Genetics. 2019;22:240–54.

8. Leskinen T, Waller K, Mutikainen S, Aaltonen S, Ronkainen PHA, et al. Effects of 32-year leisure time physical activity discordance in twin pairs on health (TWINACTIVE Study): Aims, design and results for physical fitness. Twin Res Human Genetics. 2009;12:108–17.

9. Rottensteiner M, Leskinen T, Niskanen E, Aaltonen S, Mutikainen S, et al. Physical activity, fitness, glucose homeostasis, and brain morphology in twins. Med Sci Sports Exerc. 2015;47:509–18.

10. Aaltonen S, Leskinen T, Morris T, Alen M, Kaprio J, et al. Motives and barriers to physical activity in twin pairs discordant for leisure time physical activity for 30 years. Int J Sports Med. 2012;33:157–63.

11. Aaltonen S, Rottensteiner M, Kaprio J, Kujala UM. Motives for physical activity among active and inactive persons in their mid-30s. Scand J Med Sci Sports. 2014;24:727–35.

12. Mutikainen S, Perhonen M, Alén M, Leskinen T, Karjalainen J, et al. Effects of long-term physical activity on cardiac structure and function: A twin study. J Sports Sci Med. 2009;8:533–42.

13. Leskinen T, Sipilä S, Alen M, Cheng S, Pietiläinen KH, et al. Leisure time physical activity and high-risk fat: A longitudinal population-based twin study. Int J Obesity. 2009;33:1211–8.

14. Rottensteiner M, Leskinen T, Järvelä-Reijonen E, Väisänen K, Aaltonen S, et al. Leisure-time physical activity and intra-abdominal fat in young adulthood: A monozygotic co-twin control study. Obesity. 2016;24:1185–91.

15. Rintala M, Lyytikäinen A, Leskinen T, Alen M, Pietiläinen KH, et al. Leisure time physical activity and nutrition: a twin study. Public Health Nutr. 2011;14:846–52.

16. Kujala UM, Mäkinen V-P, Heinonen I, Soininen P, Kangas AJ, et al. Long-term leisure-time physical activity and serum metabolome. Circulation. 2013;127:340–8.

17. Wilkins JT, Seckler HS. HDL modification: recent developments and their relevance to atherosclerotic cardiovascular disease. Curr Opin Lipidol. 2019;30:24–9.

18. Leskinen T, Rinnankoski-Tuikka R, Rintala M, Seppänen-Laakso T, Pöllänen E, et al. Differences in muscle and adipose tissue gene expression and cardio-metabolic risk factors in the members of physical activity discordant twin pairs. PLoS One. 2010;5:e12609.

19. Felig P, Marliss E, Cahill GF Jr. Plasma amino acid levels and insulin secretion in obesity. New Engl J Med. 1969;281:811–16.

20. Pietiläinen KH, Naukkarinen J, Rissanen A, Saharinen J, Ellonen P, et al. Global transcript profiles of fat in monozygotic twins discordant for BMI: pathways behind acquired obesity. PLoS Med. 2008;5:e51.

21. Leskinen T, Usenius J-P, Alen M, Kainulainen H, Kaprio J, et al. Leisure time physical activity and artery lumen diameters: A monozygotic co-twin control study. Scand J Med Sci Sports. 2011;21:e208–14.

22. Ma H, Leskinen T, Alen M, Cheng S, Sipilä S, et al. Long-term leisure time physical activity and properties of bone: A twin study. J Bone Mineral Res. 2009;24:1427–33.

23. Tarkka IM, Hautasaari P, Pesonen H, Niskanen E, Rottensteiner M, et al. Long-term physical activity may modify brain structure and function: Studies in young healthy twins. J Phys Act Health. 2019;16:637–43.

24. Hautasaari P, Savic AM, Loberg O, Niskanen E, Kaprio J, et al. Somatosensory brain function and gray matter regional volumes differ according to exercise history: Evidence from monozygotic twins. Brain Topogr. 2017;30:77–86.

25. Pesonen H, Savic AM, Kujala UM, Tarkka IM. Long-term physical activity modifies automatic visual processing. Int J Sport Exerc Psychol. 2019;17:275–84.

26. Kujala UM, Kaprio J, Koskenvuo M. Diabetes in a population-based series of twin pairs discordant for leisure sedentariness. Diabetologia. 2000;43:259–63.

27. Kujala UM, Kaprio J, Sarna S, Koskenvuo M. Relationship of leisure-time physical activity and mortality. The Finnish Twin Cohort. JAMA. 1998;279:440–4.

28. Kujala UM, Kaprio J, Koskenvuo M. Modifiable risk factors as predictors of all-cause mortality: The roles of genetics and childhood environment. Am J Epidemiol. 2002;156:985–93.

29. Karvinen S, Waller K, Silvennoinen M, Koch LG, Britton SL, et al. Physical activity in adulthood: genes and mortality. Scientific Reports. 2015;5:18259.

30. Waller K, Vähä-Ypyä H, Törmäkangas T, Hautasaari P, Lindgren N, et al. Long-term leisure-time physical activity and other health habits as predictors of objectively monitored late-life physical activity – A 40-year twin study. Scientific Reports. 2018;8:9400.

31. Kujala UM, Hautasaari P, Vähä-Ypyä H, Waller K, Lindgren N, et al. Chronic diseases and objectively measured physical activity among aged individuals -a cross-sectional twin cohort study. Annals Medicine. 2019;51:78–87.

32. Waller K, Kaprio J, Kujala UM. Associations between long-term physical activity, waist circumference and weight gain: a 30-year longitudinal twin study. Int J Obesity. 2008;32:353–61.

33. Kujala UM. Summary of the effects of exercise therapy in non-communicable diseases: Clinically relevant evidence from meta-analyses of randomized controlled trials. MedRxiv. 2021.02.11.21251608.

