## Supplementary information for "Physical activity and health: Findings from Finnish monozygotic twin pairs discordant for physical activity"

### Supplementary Table 1. Selected data from monozygotic twin pairs discordant for leisure time physical activity habits, TWINACTIVE and FITFATTWIN studies

| Variable | FITFATTWIN (N=10 pairs 3+ -year discordant for LTPA, mean age 34, range 32-36 years) | | | TWINACTIVE (N=7 pairs 30+ -year discordant for LTPA; mean age 62, range 50-74 years) | | | Pooled data (N=17 pairs) | | |
|---|---|---|---|---|---|---|---|---|---|
| | Less active co-twins | More active co-twins | Intrapair difference Mean (95% CI); p-value | Less active co-twins | More active co-twins | Intrapair difference Mean (95% CI); p-value | Less active co-twins | More active co-twins | Intrapair difference Mean (95% CI); p-value |
| | Mean±SD | | | Mean±SD | | | Mean±SD | | |
| **$VO_2$peak, mlxkg$^{-1}$xmin$^{-1}$** | 37.3±3.5 | 43.6±4.2 | 6.3 (4.1 to 8.5); p=0.18x10$^{-3}$ | 27.4±5.3 | 32.2±6.0 | 4.8 (-0.9 to 10.5); p=0.083 | 33.0±6.6 | 38.6±7.6 | 5.6 (3.3 to 8.0); p=0.15x10$^{-3}$ |
| **Leisure-time physical activity** | | | | | | | | | |
| **Past 3-year-LTMET index, MET-hours/day** | 1.7±1.3 | 5.0±2.7 | 3.3 (1.9 to 4.8); p=5.5x10$^{-4}$ | | | | | | |
| **Past 30-year-LTMET index, MET-hours/day** | | | | 3.0±2.6 | 11.2±4.9 | 8.2 (3.4 to 13.0); p=5.8x10$^{-3}$ | | | |
| **12-month LTMET index, MET-hours/day** | 1.2±0.9 | 3.9±1.2 | 2.8 (2.0 to 3.5); p=1.0x10$^{-5}$ | 3.2±2.2 | 9.2 ± 3.9 | 5.9 (2.1 to 9.7); p=8.8x$^{10-3}$ | 2.0±1.8 | 6.1±3.7 | 4.1 (2.5 to 5.6); p=5.3x10$^{-5}$ |
| **Body composition** | | | | | | | | | |
| **Body height, cm** | 179.1±5.2 | 179.8±5.4 | 0.7 (-0.5 to 1.8); p=0.21 | 172.8±12.0 | 173.7±11.8 | 0.9 (0.2 to 2.1); p=0.083 | 176.5±8.9 | 177.3±8.8 | 0.8 (0.1 to 1.5); p=0.034 |
| **Body weight, kg** | 77.8±12.7 | 75.8±8.5 | -2.0 (-6.9 to 2.6); p=0.38 | 77.0±11.3 | 75.4±13.2 | -1.52 (-8.9 to 5.8); p=0.63 | 77.5±11.7 | 75.7±10.3 | -1.8 (-5.5 to 1.8); p=0.31 |
| **Body-mass index, kg/m$^2$** | 24.2±3.3 | 23.4±1.7 | -0.8 (-2.3 to 0.8); p=0.28 | 25.7 ± 1.5 | 24.8 ± 1.9 | -0.9 (-3.4 to 1.7); p=0.44 | 24.8±2.7 | 24.0±1.9 | -0.8 (-2.0 to 0.4) p=0.17 |
| **Waist circumference, cm** | 88.6±8.2 | 85.3±6.2 | -3.3 (-7.4 to 0.8); p=0.099 | 97.6±7.9 | 92.9±8.1 | -4.7 (-10.8 to 1.4); p=0.11 | 92.3±9.0 | 88.4±7.8 | -3.9 (-6.9 to -0.8); p=0.016 |
| **Fat percent, %** | 19.0±2.9 | 17.4±3.2 | -1.6 (-3.7 to 0.6;); p=0.13 | 27.9±5.6 | 22.9±4.7 | -5.0 (-11.3 to 1.4); p=0.10 | 22.6±6.0 | 19.7±4.7 | -3.0 (-5.6 to -0.6); p=0.027 |
| **Visceral adipose tissue area, cm$^2$** | 129±54 | 92±47 | -37 (-66 to -8); p=0.018 | 165±51 | 128±62 | -37 (-89 to 16); p=0.14 | 144±54 | 107±55 | -37 (-61 to -13); p=0.0045 |
| **Subcutaneous adipose tissue area, cm$^2$** | 115±36 | 99±35 | -16 (-42 to 11); p=0.21 | 185±40 | 161±26 | -25 (-63 to 13); p=0.16 | 144±51 | 125±44 | -19 (-39 to -0.2); p=0.048 |
| **Visceral per subcutaneous adipose tissue ratio** | 1.123±0.325 | 0.932 ±0.330 | -0.191 (-0.380 to -0.002); p=0.049 | 0.924±0.329 | 0.813±0.383 | -0.110 (-0.394 to 0.173); p=0.38 | 1.041±0.332 | 0.883±0.346 | -0158 (-0.229 to -0.016); p=0.032 |
| **Liver fat index, MRI signal intensity** | 7.3±3.9 | 7.3±4.0 | -0.0 (-2.2 to 2.1); p=0.98 | 26.1±21.6 | 6.0±8.2 | -20.1 (-34.0 to 6.2); p=0.012 | 15.1±16.6 | 6.8±5.9 | -8.3 (-15.4 to -1.1); p=0.026 |
| **Lipoproteins** | | | | | | | | | |
| **Apo-B, g/L** | 1.109±0.143 | 0.926 ±0.186 | -0.182 (-0.402 to 0.037); p=0.089 | 0.796±0.152 | 0.745±0.193 | -0.051 (-0.119 to 0.017); p=0.13 | 0.925±0.214 | 0.820±0.206 | -0.105 (-0.195 to -0.015); p=0.025 |
| **Apo-A1, g/L** | 1.806±0.283 | 1.824 ±0.271 | 0.018 (-0.057 to 0.093); p=0.58 | 1.373±0.240 | 1.412±0.235 | 0.039 (-0.079 to 0.158); p=0.47 | 1.551±0.333 | 1.582±0.320 | 0.033 (-0.039 to 0.100); p=0.36 |
| **ApoB:ApoA1 ratio** | 0.628±0.167 | 0.522 ±0.167 | -0.105 (-0.246 to 0.035); p=0.12 | 0.593±0.137 | 0.533±0.125 | -0.060 (-0.104 to -0.016); p=0.013 | 0.607±0.133 | 0.529±0.034 | -0.079 (-0.133 to -0.024); p=0.0077 |

| | | | | | | | | | |
|---|---|---|---|---|---|---|---|---|---|
| **HDL cholesterol, mmol/L** | 1.664±0.392 | 1.815 ±0.447 | 0.151 (-0.034 to 0.336); p=0.093 | 1.261±0.319 | 1.347±0.313 | 0.086 (-0.060 to 0.233); p=0.22 | 1.427±0.096 | 1.540±0.105 | 0.113 (0.011 to 0.216); p=0.033 |
| **$HDL_2$ cholesterol, mmol/L** | 1.117±0.369 | 1.291 ±0.460 | 0.174 (-0.035 to 0.382); p=0.088 | 0.795±0.276 | 0.873±0.295 | 0.078 (-0.045 to 0.202); p=0.19 | 0.928±0.347 | 1.045±0.416 | 0.118 (0.017 to 0.218); p=0.025 |
| **HDL diameter, nm** | 9.829±0.221 | 9.929 ±0.254 | 0.100 (-0.015 to 0.215); p=0.077 | 9.648±0.215 | 9.761±0.210 | 0.113 (0.044 to 0.181); p=0.00487 | 9.722±0.230 | 9.830±0.237 | 0.108 (0.054 to 0.162); p=6.5x$10^{-4}$ |
| **Very large HDL particles, mmol/L** | 2.354±1.686 | 3.144 ±1.750 | 0.760 (-0.031 to 1.833); p=0.13 | 1.868±1.078 | 2.458±1.456 | 0.586 (-0.001 to 1.182); p=0.052 | 2.068x$10^{-4}$ ±0.1334x$10^{-4}$ | 2.728x$10^{-4}$ ±1.566x$10^{-4}$ | 0.660x$10^{-4}$ (0.171x$10^{-4}$ to 1.148x$10^{-4}$); p=0.011 |
| **Large HDL particles, mmol/L** | 10.472±4.206 | 12.659 ±5.336 | 2.187 (-0.001 to 4.386); p=0.051 | 6.300±3.706 | 7.735±3.737 | 1.435 (0.336 to 2.533); p=0.016 | 8.018 x$10^{-4}$ ±4.340 x$10^{-4}$ | 9.762x$10^{-4}$ ±4.977x$10^{-4}$ | 1.744x$10^{-4}$ (0.769x$10^{-4}$ to 2.719x$10^{-4}$); p=0.0016 |
| **Medium HDL particles, mmol/L** | 22.035±3.241 | 21.907 ±3.900 | -0.013 (-0.252 to 2.266); p=0.90 | 15.510±3.498 | 15.522±2.168 | 0.012 (-0.181 to 1.837); p=0.99 | 18.197x$^{10-4}$ ±4.667x$^{10-4}$ | 18.151x$^{10-4}$ ±4.341x$^{10-4}$ | -0.0453x$^{10-4}$ (-1.323x$^{10-4}$ to 1.232 x$^{10-4}$); p=0.94 |
| **Small HDL particles, mmol/L** | 49.635±2.271 | 48.794 ±3.197 | -0.084 (-0.299 to 1.309); p=0.38 | 44.106±2.272 | 43.115±2.581 | -0.099 (-0.355 to 1.566); p=0.40 | 46.383x$^{10-4}$ ±3.564x$^{10-4}$ | 45.453x$^{10-4}$ ±3.925 x$^{10-4}$ | -0.930x$^{10-4}$ (-2.491x$^{10-4}$ to 0.632x$^{10-4}$); p=0.23 |

Abbreviations: CI, confidence interval; LTMET-hours/day, leisure-time metabolic equivalent for physical activity during leisure-time and the commute to and from work indicating daily leisure-time physical activity volume; MRI, magnetic resonance imaging; ApoB, apolipoprotein B; ApoA1, apolipoprotein A1; HDL, high density lipoprotein.

**Supplement 2. Links to PhD theses (Sports and Exercise Medicine, University of Jyväskylä) which include results from leisure-time physical activity discordant twin pairs**


1. Waller Katja. Leisure-time physical activity, weight gain and health. A prospective follow-up in twins. University of Jyväskylä, 2011. **http://urn.fi/URN:ISBN:978-951-39-4450-6**

2. Leskinen Tuija. Long-term leisure-time physical activity vs. inactivity, physical fitness, body-composition and metabolic health characteristics: a co-twin control study. University of Jyväskylä, 2013. **http://urn.fi/URN:ISBN:978-951-39-5209-9**

3. Rottensteiner Mirva. Leisure-time physical activity habits and abdominal adiposity in young adulthood. Twin cohort and co-twin control studies. University of Jyväskylä, 2018. **http://urn.fi/URN:ISBN:978-951-39-7472-5**

4. Aaltonen Sari. Leisure-time physical activity in a Finnish twin study. Genetic and environmental influences as determinants and motives as correlates. University of Jyväskylä, 2013. **http://urn.fi/URN:ISBN:978-951-39-5326-3**

5. Mutikainen Sara. Genetic and environmental effects on resting electrocardiography and the association between electrocardiography and physical activity, walking endurance and mortality in older people. University of Jyväskylä, 2010. **http://urn.fi/URN:ISBN:978-951-39-4153-6**

6. Ma Hongqiang. Adaptation of bone to physical activity and diet-induced obesity. University of Jyväskylä, 2011. **http://urn.fi/URN:ISBN:978-951-39-4401-8**

7. Föhr Tiina. The relationship between leisure-time physical activity and stress on workdays with special reference to heart rate variability analyses. University of Jyväskylä, 2016. **http://urn.fi/URN:ISBN:978-951-39-6794-9**

8. Hautasaari Pekka. Exercise effects on early cortical somatosensory and nociceptive processing in the human brain. University of Jyväskylä, 2019. **http://urn.fi/URN:ISBN:978-951-39-7949-2**

9. Karvinen Sira. Lifespan and skeletal muscle properties. The effects of genetic background, physical activity and ageing. University of Jyväskylä, 2016. **http://jyx.jyu.fi/handle/123456789/49353**
